# Diagnostic delay of Myositis: protocol for an integrated systematic review

**DOI:** 10.1101/2022.04.27.22274397

**Authors:** Tergel Namsrai, Jane Desborough, Anita Chalmers, Christine Lowe, Matthew Cook, Christine Phillips, Anne Parkinson

## Abstract

Idiopathic inflammatory myopathies (IIM) described as “inflammatory myositis”, are a heterogeneous group of rare muscular autoimmune diseases characterized by skeletal muscle inflammation. Its complex characteristics with lack of accurate diagnostic tests, unified classification system, and comprehensive widely used diagnostic criteria could lead to diagnostic delay. This study will review diagnostic delay in Myositis and provide an overview and clearer insight of patients’ experiences, causes and consequences of diagnostic delay in Myositis.

**Methods and analysis:** The literature source will be a systematic search of PubMed/Medline, Scopus, ProQuest, and sources of grey literature, conducted from database inception to 15^th^ of December 2021 without restrictions on publication date. All study types (qualitative and quantitative) except review articles, examining diagnostic delay, incorrect diagnosis, missed diagnosis or slow diagnosis of all types of myositis in all ages will be included. Evidence of patients’ experiences associated with diagnostic delay will also be examined. Studies in languages other than English, German and Indonesian will be excluded. Outcomes will be diagnostic delay time, patients’ experiences, and causes and consequences associated with diagnostic delay in Myositis. Two review authors will independently screen the titles and abstracts of search results against the inclusion criteria. The Mixed Methods Appraisal Tool (MMAT) will be used to appraise selected studies. Two independent authors will extract data using a pre-piloted data extraction tool. If sufficient quantitative data is available, a meta-analysis will be conducted along with subgroup analysis including pooled diagnostic delay in each type of Myositis. Qualitative data will be analysed in line with meta-aggregation methods. If data is insufficient, a narrative synthesis will be conducted.

**Ethics and dissemination:** As a systematic review, ethical approval was not required. Findings of the study will be disseminated through publications in peer-reviewed journals, conferences, and symposia.

**PROSPERO Registration number:** CRD42022289830

**Strengths and limitations of this study:**

1. The protocol was developed in accordance with Preferred Reporting Items for Systematic Review and Meta-Analysis Protocols guidelines and the Cochrane Handbook for Systematic Reviews.
2. Examination of both quantitative and qualitative literature will enable insight into the causes and consequences, and patients’ experiences associated with diagnostic delay of Myositis.
3. In some case studies, it will be necessary to manually calculate diagnostic delay according to the date of symptom onset and the date of diagnosis.
4. The primary limitation of this study is that it will not capture evidence that is not published, or from situations when an accurate diagnosis of Myositis is not made.
5. The review will be limited to English, German, and Indonesian studies.

## Introduction

Idiopathic inflammatory myopathies (IIM) commonly described as “inflammatory myositis”, are a heterogenous group of rare muscular diseases characterized as skeletal muscle inflammation and other extra muscular features such as skin manifestations ^1^.

There are several subtypes of IIM including dermatomyositis (DM), polymyositis (PM), inclusion body myositis (IBM) and other specified idiopathic myositis (i.e. immune-mediated necrotizing myopathy (IMNM), juvenile myositis (JM), juvenile dermatomyositis (JDM), amyopathic dermatomyositis (AMD) and anti-synthetase syndrome (ASS)), and unspecified idiopathic inflammatory myositis ^2^.

IIM is characterized as a rare disease as its prevalence is relatively low compared to other disorders. A recent systematic review of 16 articles reported an overall estimated incidence rate of 78 cases/100,000 per year for IIM ^3^.

However, IIM has broad clinical characteristic features involving both muscular and extra-muscular systems with acute or progressive onset. In addition to general muscle features it can present with dysphagia (39%), lung involvement causing interstitial lung disease (ILD) (30%), malignancy (13%), and cardiac disease (9%) ^4^.

There has been significant promising progress on Myositis Specific Autoantibodies in the last decade. The presence of these antibodies assists the suspected diagnosis of IIM ^5^. Additionally, MRI imaging can reveal specific changes in the involved muscle and therefore aids the diagnostic process of IIM ^6^. However, there is a lack of conclusive diagnostic tests and commonly used comprehensive diagnostic criteria. The most widely used criteria is Bohan and Peter’s criteria which recognizes PM and DM as IIM ^7^. Later, Dalakas introduced different criteria which take into account AMD ^8^. However, these two criteria both still exclude IBM as an individual type of IIM.

In 2017, the European League Against Rheumatism (EULAR) and the American College of Rheumatism (ACR), developed diagnostic and classification criteria based on the data from 976 IIM cases and 624 comparators ^9^. The EULAR/ARC criteria permit specialists to differentiate between all possible IIM subgroups that are not mentioned in previously used criteria, including JM, JDM and IMNM.

Due to the low prevalence, broad range of clinical features, lack of conclusive diagnostic testing and comprehensive globally accepted criteria, timely diagnosis of IIM can be challenging and result in significant diagnostic delays. Some studies have reported diagnostic delay of 4-5.6 years in cases of IBM ^10 11^. However, studies examining the overall diagnostic delay, factors associated with diagnostic delay, and people’s experience of diagnostic delay in IIM are scarce. Further studies are crucial for gaining clearer insight into diagnostic delays. This will inform future studies, interventions, tools, and health policies directed at enhancing diagnostic efficiency and patient experience of Myositis.

## Objective

The aim of this integrated systematic review is to review the evidence regarding diagnostic delay in Myositis. To this end, the systematic review will aim to answer the following research questions:

RQ1. What are the causes and consequences of diagnostic delay of myositis?

RQ2. What evidence is there about patients’ experience of Myositis’ diagnostic delay?

## Methods and Analysis

### Protocol development

This study protocol is based on the Preferred Reporting Items for Systematic Review and Meta-Analysis Protocols (PRISMA-P) and the Cochrane Handbook for Systematic Reviews ^12 13^.

### Search strategy

The search strategy was developed to ensure reproducibility and increase transparency following the PRISMA-P checklist ^12^. Research questions and search terms were developed using the PICOS tool (Population/Intervention/Comparison/Outcomes/Study Design) to enhance the scientific literature by ensuring reliability and homogeneity of search results ^14^. The study is registered with PROSPERO (CRD42022289830). The primary source of literature will be a systematic search of multiple electronic databases (from inception onwards): PubMed/Medline, Scopus, and ProQuest. Sources of grey literature will also be searched. The grey literature search will be conducted through Open Access Theses and Dissertation (https://oatd.org/), ProQuest thesis and dissertations, The National Library of Australia, and The Myositis Association Australia website (https://myositis.org.au/). Additionally, reference lists of selected studies and review articles will be searched. All settings and study design will be considered.

Search terms were developed in collaboration with research team members (TN, AP, JD). Search terms were combined using Boolean operators “AND” and “OR”. A preliminary exploratory search on PUBMED/MEDLINE was undertaken (15^th^ October 2021) as shown in Table 1 to inform the final search strategy and determine outcomes. This search strategy was updated and peer reviewed (MC, CP) using the PRESS checklist ^15^. The final search terms included Myositis AND (“delay in diagnosis” OR “diagnostic delay” OR “misdiagnosis” OR “time to diagnosis” OR “incorrect diagnosis” OR “missed diagnosis” OR “delayed diagnosis”) without restrictions on study type, date, and language.

**Table 1.** Search string conducted on PUBMED/MEDLINE

| Table 1. Search string conducted on PUBMED/MEDLINE |  |  |
| --- | --- | --- |
| Search number | Query | Search Details |
| 1 | myositis[Title/Abstract] | "myositis"[Title/Abstract] |
| 2 | "delay in diagnosis"[Title/Abstract] | "delay in diagnosis"[Title/Abstract] |
| 3 | "diagnostic delay"[Title/Abstract] | "diagnostic delay"[Title/Abstract] |
| 4 | "misdiagnosis"[Title/Abstract] | "misdiagnosis"[Title/Abstract] |
| 5 | "time to diagnosis"[Title/Abstract] | "time to diagnosis"[Title/Abstract] |
| 6 | "incorrect diagnosis"[Title/Abstract] | "incorrect diagnosis"[Title/Abstract] |
| 7 | "missed diagnosis"[Title/Abstract] | "missed diagnosis"[Title/Abstract] |
| 8 | "delayed diagnosis"[Title/Abstract] | "delayed diagnosis"[Title/Abstract] |
| 9 | "slow diagnosis"[Title/Abstract] | "slow diagnosis"[Title/Abstract] |
| 10 | #2 OR #3 OR #4 OR #5 OR #6 OR #7 OR #8 OR #9 | "delay in diagnosis"[Title/Abstract] OR "diagnostic delay"[Title/Abstract] OR "misdiagnosis"[Title/Abstract] OR "time to diagnosis"[Title/Abstract] OR "incorrect diagnosis"[Title/Abstract] OR "missed diagnosis"[Title/Abstract] OR "delayed diagnosis"[Title/Abstract] |
| 11 | #1 AND #10 | "myositis"[Title/Abstract] AND ("delay in diagnosis"[Title/Abstract] OR "diagnostic delay"[Title/Abstract] OR "misdiagnosis"[Title/Abstract] OR "time to diagnosis"[Title/Abstract] OR "incorrect diagnosis"[Title/Abstract] OR "missed diagnosis"[Title/Abstract] OR "delayed diagnosis"[Title/Abstract]) |

The final search string used for the literature search conducted on 9^th^ of December is included in Supplementary Table 1.

### Study selection

The literature search results will be imported to Covidence, an internet-based software that facilitates collaboration between reviewers and ensures independent review of the literature ^16^.

Studies will be selected according to the pre-developed PICOS eligibility criteria outlined in Table 2. We will include all types of studies including both qualitative and quantitative, examining diagnostic delay, incorrect diagnosis, missed diagnosis or slow diagnosis of all types of myositis including dermatomyositis, polymyositis, necrotizing myositis, juvenile dermatomyositis, inclusion body myositis, mixed connective tissue diseases, overlap myositis, interstitial myositis, orbital myositis and antisynthetase syndromes in all age groups. Evidence of patients’ experiences associated with diagnostic delay will also be examined. There will be no comparison group given the nature of the study. No setting or publication date restrictions will be imposed. However, review studies and studies in languages other than English, German and Indonesian will be excluded.

**Table 2.**
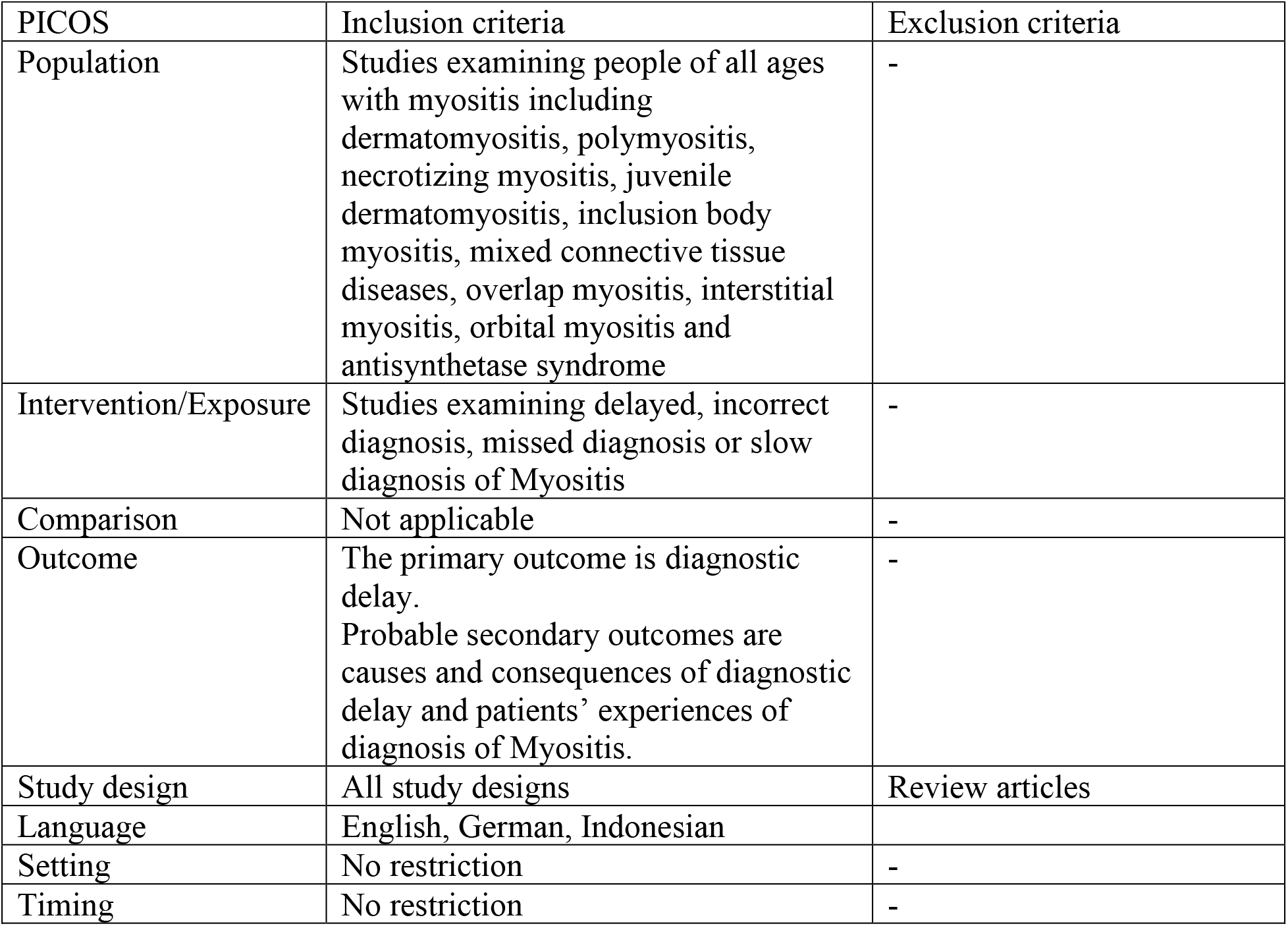
Inclusion and exclusion criteria

First, two review authors (TN and AP) will independently screen the titles and abstract of the literature search results against the pre-developed inclusion criteria. Any conflict in the title and abstract screening process will be discussed among the review team and will be resolved by a third reviewer (JD).

Full reports for all studies that meet the inclusion criteria or where there is any uncertainty will be obtained. Review authors (TN and AP) will then screen full text reports according to the inclusion criteria. Any conflicts will be resolved by a third reviewer (JD). The reasons for excluding studies will be recorded. Authors will not be blinded to the study types, journals, and authors during this process.

## Data extraction

After the study selection process is complete, a data extraction tool will be designed, peer reviewed and piloted. In the piloting process, two independent reviewers (TN and AP) will extract data independently and in duplicate from five studies each and compare their results to establish agreement and validity of the data extraction tool.

Data items to be extracted include:

1. Identification of the study (journal, authors, year, citation, research center/university/hospital/organization, conflict of interest, funding/sponsorship),
2. Methods (study aim, study design, participant demographics, recruitment process, inclusion, exclusion criteria, statistical analysis),
3. Main findings (exposure details, diagnostic delays, causes and consequences of delay, patients’ experience, and other relevant outcomes).

In cases of missing diagnostic delay, date of symptom onset and date of diagnosis will be used to calculate diagnostic delay. The correct diagnosis is defined as diagnosis of Myositis regardless of the type. Any disagreements will be resolved through discussion and conflicts resolved by a third reviewer (JD). We will contact study authors to resolve any uncertainties about extracted data.

The primary outcome of the review is diagnostic delay time (time from symptom onset to correct diagnosis) in people living with Myositis. Additional secondary outcomes include patient’s experiences, causes and consequences of diagnostic delay in Myositis.

### Quality appraisal

The selected studies will be assessed for methodological quality or risk of bias using the Mixed Methods Appraisal Tool (MMAT) designed to critically appraise mixed method studies included in systematic reviews ^17^. Two independent review authors (TN and AP) will conduct the quality appraisal. Any conflicts will be resolved with discussion and a third reviewer’s vote (JD).

### Data synthesis and meta-analysis

A systematic narrative synthesis will be undertaken to explore the findings of included studies in relation to time from symptom onset to diagnosis, and people’s experiences related to delayed diagnosis in line with guidance from the Centre for Reviews and Dissemination ^18^.

If extracted quantitative data are homogenous, a meta-analysis will be conducted using a random-effects model along with subgroup analysis including pooled diagnostic delay in each type of Myositis (dermatomyositis, polymyositis, juvenile dermatomyositis, inclusion body myositis, antisynthetase syndrome and others). Extracted qualitative data will be meta-synthesized using meta-aggregation. In line with meta-aggregation methods, findings (processed data) from qualitative studies will be extracted and aggregated into a single set of categories, which will then be further aggregated and synthesised into a set of statements that are meaningful for clinical practice.

Further methods and stages of meta-analysis will be discussed if collected data is quantitively synthesizable. The findings from the quantitative and qualitative studies will be reported separately; however, the discussion will be integrative of both.

### Quality of evidence

If a meta-analysis is conducted, the quality/certainty of evidence for all quantitative outcomes will be judged using the Grading of Recommendations Assessment, Development and Evaluation (GRADE) working group methodology ^19^. Certainty of the body of evidence will be assessed across domains of risk of bias, consistency of effect, imprecision, indirectness, and publication bias. The certainty will be reported in four levels: high, moderate, and very low.

### Amendments

In the event of protocol amendments prior to study commencement, date, explanation, and rationale of the amendment will be described in the final protocol. The record will be in tabular format as recommended by the Cochrane Collaboration^12^.

### Patient and public involvement

We follow a co-production approach in all our research. The research team includes two members with Myositis, an immunologist, a general practitioner (GP), and a registered nurse.

## Data Availability

All data produced in the present study are available upon reasonable request to the authors

## Ethics and dissemination

As a systematic review, no human will be involved or participated in the study, with no necessity for ethical approval. Findings of the study will be disseminated through publications in peer-reviewed journals, conferences, and symposia.

## Author contributions

All authors were equally involved in the study.

JD, AC, CL, MC, CP, and AP developed and designed the study concept. TN prepared the study protocol and drafted the manuscript. JD and AP supervised the procedure of developing the study protocol. MC, CP, AC, and CL reviewed the study protocol.

