## Supplementary file for "Diagnostic delay of Myositis: protocol for an integrated systematic review"

Supplementary table 1. Myositis literature search

Online Database search

1. MEDLINE/PUBMED search

09-Dec-21

| Search number | Query | Search Details | Results | Selected results for screening |
|---|---|---|---|---|
| 11 | #1 AND #10 | "myositis"[Title/Abstract] AND ("delay in diagnosis"[Title/Abstract] OR "diagnostic delay"[Title/Abstract] OR "misdiagnosis"[Title/Abstract] OR "time to diagnosis"[Title/Abstract] OR "incorrect diagnosis"[Title/Abstract] OR "missed diagnosis"[Title/Abstract] OR "delayed diagnosis"[Title/Abstract] OR "slow diagnosis"[Title/Abstract]) | 96 | |
| 10 | #2 OR #3 OR #4 OR #5 OR #6 OR #7 OR #8 OR #9 | "delay in diagnosis"[Title/Abstract] OR "diagnostic delay"[Title/Abstract] OR "misdiagnosis"[Title/Abstract] OR "time to diagnosis"[Title/Abstract] OR "incorrect diagnosis"[Title/Abstract] OR "missed diagnosis"[Title/Abstract] OR "delayed diagnosis"[Title/Abstract] | 38,185 | |
| 9 | "slow diagnosis"[Title/Abstract] | "slow diagnosis"[Title/Abstract] | 8 | |
| 8 | "delayed diagnosis"[Title/Abstract] | "delayed diagnosis"[Title/Abstract] | 8,589 | |
| 7 | "missed diagnosis"[Title/Abstract] | "missed diagnosis"[Title/Abstract] | 2,399 | |
| 6 | "incorrect diagnosis"[Title/Abstract] | "incorrect diagnosis"[Title/Abstract] | 1,299 | |
| 5 | "time to diagnosis"[Title/Abstract] | "time to diagnosis"[Title/Abstract] | 2,661 | |
| 4 | "misdiagnosis"[Title/Abstract] | "misdiagnosis"[Title/Abstract] | 16,610 | |
| 3 | "diagnostic delay"[Title/Abstract] | "diagnostic delay"[Title/Abstract] | 3,203 | |
| 2 | "delay in diagnosis"[Title/Abstract] | "delay in diagnosis"[Title/Abstract] | 5,967 | |
| 1 | myositis[Title/Abstract] | "myositis"[Title/Abstract] | 11,085 | |

2. ProQuest search

09-Dec-21

| Search number | Query | Search details | Results | Selected results for screening |
|---|---|---|---|---|
| 1 | 1 | ab("myositis") AND ab("delay in diagnosis" OR "delayed diagnosis" OR "diagnostic delay" OR "time to diagnosis" OR "misdiagnosis" OR "missed diagnosis" OR "incorrect diagnosis" OR “slow diagnosis”) | 35 | |

3. Scopus search

09-Dec-21

| Search number | Query | Search details | Results | Selected results for screening |
|---|---|---|---|---|
| 1 | 1 | TITLE-ABS ( "myositis" ) AND TITLE-ABS ( "delay in diagnosis" ) OR TITLE-ABS ( "delayed diagnosis" ) OR TITLE-ABS ( "diagnostic | 113 | |

| | | | | |
|---|---|---|---|---|
| | | delay" ) OR TITLE-ABS ( "time to diagnosis" ) OR TITLE-ABS ( "misdiagnosis" ) OR TITLE-ABS ( "missed diagnosis" ) OR TITLE-ABS ( "incorrect diagnosis" ) OR TITLE-ABS ( "slow diagnosis" ) | | |
| Grey literature search | | | | |
| 1. ProQuest dissertations, thesis, reports and conference presentations | | | | |
| Search number | Query | Search details | Results | Selected for screening |
| 1 | 1 | ab('myositis) AND ab(diagnos*) | 38 | 14 |
| 2. OATD search | | | | |
| Search number | Query | Search details | Results | Selected for screening |
| 1 | 1 | abstract:(myositis) AND abstract:(diagnos*) | 46 | 4 |
| 4. Myositis Australia website = 0 found | | | | |
| 5. Back ward referencing | | | | |
| Review article name | References found | Selected for screening | | |
| Article name | References found | Selected for screening | | |

### Diagnostic delay of Myositis: a protocol of an integrated systematic review

#### Section 1: Administration Information

##### Item 1. Title

Diagnostic delay of Myositis: a protocol of an integrated systematic review

##### Item 2. Registration

This systematic review will be registered with PROSPERO.

##### Item 3. Authors

Tergel Namsrai MD, MSc[1]

Jane Desborough RN, RM, MPH, PhD[1]*

Anita Chalmers OAM[1,2]

Christine Lowe[1,2]

Matthew Cook MBBS, PhD, FRACP, FRCPA, FFSc(RCPA)[3]

Christine Phillips MBBS, BMedSc, MA, MPH, DipEd, FRACGP, MD, AM[4]

Anne Parkinson BA (Hons), AFHEA, PhD[1]

##### Item 3a. Affiliations

1. National Centre for Epidemiology and Population Health, Australian National University, Canberra, Australia

2. The Myositis Association- Australia Inc

3. John Curtin School of Medical Research, Australian National University, Canberra, Australia

4. Australian National University Medical School, Canberra, Australia

*Correspondence

Jane Desborough, National Centre for Epidemiology and Population Health, Australian National University, 63, Eggleston Road, Acton ACT, 2601, Australia

##### Item 3b. Contributions

TN drafted the review protocol. All authors will contribute to the study. TN and AP are the primary reviewers. JD is the third reviewer guarantor of the study.

##### Item 4. Amendments

In the event of protocol amendments, date, explanation, and rationale of the amendment will be described in this section. The record will be in tabular format as shown below.

| Table1. Record of Amendments | | | | |
|---|---|---|---|---|
| Date | Section | Original protocol | Revised protocol | Rationale |
| 09/12/2021 | Appendix 1. Search string | "myositis"[Title/Abstract] AND ("delay in diagnosis"[Title/Abstract] OR "diagnostic delay"[Title/Abstract] OR "misdiagnosis"[Title/Abstract] OR "time to diagnosis"[Title/Abstract] OR "incorrect diagnosis"[Title/Abstract] OR "missed diagnosis"[Title/Abstract] OR "delayed diagnosis"[Title/Abstract]) | "myositis"[Title/Abstract] AND ("delay in diagnosis"[Title/Abstract] OR "diagnostic delay"[Title/Abstract] OR "misdiagnosis"[Title/Abstract] OR "time to diagnosis"[Title/Abstract] OR "incorrect diagnosis"[Title/Abstract] OR "missed diagnosis"[Title/Abstract] OR "delayed diagnosis"[Title/Abstract]) OR “slow diagnosis”[Title/Abstract]) | After peer review “slow diagnosis” was added to the search terms. The search string has changed accordingly. |

#### Item 5. Support

##### Item 5a. Sources

This integrated systematic review is part of the “Missed opportunities in clinical practice: Tools to enhance healthcare providers’ awareness and diagnosis of rare diseases in Australia” project funded by the Commonwealth represented by the Department of Health Australia (Grant ID 4-G5ZN0T7).

##### Item 5b and 5c. Sponsor name and its role

The Commonwealth of Australia represented by the Department of Health has provided a grant for the “Missed opportunities in clinical practice: Tools to enhance healthcare providers’ awareness and diagnosis of rare diseases in Australia” project which includes this review.

### Section 2: Introduction

#### Item 6. Rationale

Diagnostic delay of Myositis: a protocol of an integrated systematic review

Idiopathic inflammatory myopathies (IIM) commonly described as "inflammatory myositis", are a heterogenous group of rare muscular autoimmune diseases of muscle inflammation initially presenting with asymmetric distal muscle weaknesses (finger flexor and knee flexor muscles) progressing over time to larger muscle weaknesses (gluteus, quadriceps and throat muscles) and other extra muscular features such as skin manifestations (Malik et al., 2016).

There are several subtypes of IIM including dermatomyositis (DM), polymyositis (PM), Inclusion body myositis (IBM) and other specified idiopathic myositis (i.e. immune-mediated necrotizing myopathy (IMNM), juvenile myositis (JM), juvenile dermatomyositis (JDM), amyopathic dermatomyositis (AMD) and anti-synthetase syndrome (ASS)), and unspecified idiopathic inflammatory myositis (World Health Organization, 2019).

IIM is characterized as a rare disease as its prevalence is relatively low compared to other disorders. A recent systematic review of 16 articles reported an overall estimated incidence rate of 78 cases/100,000 per year for IIM (Meyer et al., 2014).

However, IIM has broad clinical characteristic features involving both muscular and extra-muscular systems with acute or progressive onset. In addition to general muscle features it can present with dysphagia (39%), lung involvement causing interstitial lung disease (ILD) (30%), malignancy (13%), and cardiac disease (9%) (Lilleker et al., 2017).

There has been significant and promising progress on Myositis Specific Autoantibodies in the last decade. The presence of these antibodies assists the suspected diagnosis of IIM (Satoh et al., 2017). Additionally, MRI imaging can reveal specific changes in the involved muscle and therefore aids the diagnostic process of IIM (Maurer and Walker, 2015). However, there is a lack of any conclusive diagnostic test and commonly used comprehensive diagnostic criteria. The most widely used criteria is Bohan and Peter's criteria which recognizes PM and DM as IIM (Bohan and Peter, 1975). Later, Dalakas introduced different criteria which take into account AMD (Dalakas and Hohlfeld, 2003). However, these two criteria both still exclude IBM as an individual type of IIM.

In 2018, the European League Against Rheumatism (EULAR) and the American College of Rheumatism (ACR), developed diagnostic and classification criteria based on the data from 976 IIM cases and 624 comparators (Bottai et al., 2017). The EUCLAR/ARC criteria permit specialists to differentiate between all possible IIM subgroups that are not mentioned in previously used criteria, including JM, JDM and IMNM.

Due to the low prevalence, broad range of clinical features, lack of conclusive diagnostic testing and comprehensive globally accepted criteria, timely diagnosis of IIM can be challenging and result in significant diagnostic delays. Some studies reported diagnostic delay of 4-5.6 years in cases of IBM. However, studies examining the overall diagnostic delay, contributing factors, and people's experience in IIM are scarce. Further studies are crucial for gaining clearer insight into diagnostic delays. This will inform future studies, interventions, tools, and health policies directed at enhancing diagnostic efficiency and patient experience of Myositis.

#### Item 7. Objectives

The aim of this integrated systematic review is to review the evidence regarding diagnostic delay in Myositis. To this end, the review will answer the following questions:

1. What are the causes and consequences of diagnostic delay of Myositis?
2. What evidence is there about patients' experience of diagnostic delay of Myositis?

### Section 3: Methods

#### Item 8. Eligibility criteria

The studies will be selected according to the eligibility criteria developed using the PICOS tool (Methley et al., 2014)

Inclusion criteria:

1) Participants

   We will include all studies examining people of all ages with myositis including dermatomyositis, polymyositis, necrotizing myositis, juvenile dermatomyositis, inclusion body myositis, mixed connective tissue diseases, overlap myositis, interstitial myositis, orbital myositis and antisynthetase syndrome.

2) Exposure

   We will include all studies examining delayed, incorrect, or missed diagnosis of Myositis (outlined above in section 1 Participants).

3) Comparison or control group

   Given the aim of the study we will not include a control group.

4) Outcome of interest

   The main outcomes of interest are time to diagnosis, factors associated with diagnostic delay and patients' experiences of diagnosis of Myositis.

   We will include quantitative studies with adequately reported data (the actual words of the participant or the field notes of observers) as well as findings (the results of the researcher's analysis and interpretation).

   We will include qualitative studies with patients' experience of diagnostic delay for Myositis.

5) Timing

   There will be no restriction in timing of the studies.

6) Setting

   There will be no restriction in settings.

7) Study design

We will include all types of study design such as observational studies, clinical trials, case-reports, and qualitative studies, except for review articles. However, the reference lists of review articles will be hand searched for relevant papers.

8) Language

We will include studies published in English, Indonesian and German.

#### Item 9. Information sources

Electronic database and grey literature searches will be conducted.

1) Electronic database searches: PUBMED/MEDLINE, Scopus, and ProQuest.
2) Other methods to identify relevant literature: grey literature will be searched using Google Scholar.

The search strategy will be developed using the PICOS method as recommended in the Cochrane systematic review handbook (Methley et al., 2014).

#### Item 10. Search strategy

The search strategy was developed to ensure reproducibility and increase transparency following the PRISMA-P checklist (Moher et al., 2015). Research questions and search terms were developed using the PICOS tool (Population/Intervention/Comparison/Outcomes/Study Design) to enhance the scientific literature by ensuring reliability and homogeneity of search results (Methley et al., 2014).

The primary source of literature will be a systematic search of multiple electronic databases (from inception onwards): PubMed/Medline, Scopus, and ProQuest. Sources of grey literature will also be searched. A search of the grey literature will be conducted through Open Access Theses and Dissertation (https://oatd.org/ ), ProQuest thesis and dissertations, The National Library of Australia, and The Myositis Association Australia website (https://myositis.org.au/ ). Additionally, reference lists of selected studies and review articles will be searched. All settings and study design will be considered.

Search terms were developed in collaboration with research team members and peer reviewed (TN, AP, JD, MC, CP) using the PRESS checklist (McGowan et al., 2016). Search terms were combined using Boolean operators "AND" and "OR". Preliminary exploratory searches of the literature were undertaken (15 October 2021) to inform the final search strategy and determine outcomes. The final search strategy that was developed and used on PUBMED/MEDLINE database is shown in Appendix 1.

#### Item 11. Study records

##### Item 11a. Data management

The literature search results will be imported to Covidence, an internet-based software that facilitates collaboration between reviewers and ensures independent review of the literature (Veritas Health Innovation)

##### Item 11b. Selection process

Two review authors will independently screen the titles and abstracts of literature identified in the search against the pre-developed inclusion criteria (TN and AP). Any conflict in the title and abstract screening process will be discussed among the review team and will be resolved by a third reviewer (JD). Full reports for all studies that meet the inclusion criteria or where there is any uncertainty will be obtained. Review authors will then screen full text reports according to the inclusion criteria. The reasons for excluding studies will be recorded. Authors will not be blinded to the study types, journals, and authors during this process.

##### Item 11c. Data collection

After the study selection process is complete, a data extraction tool will be designed, peer reviewed and piloted. In the piloting process, two independent reviewers will extract data independently and in duplicate from five studies each and compare their results to establish agreement and validity of the data extraction tool. Any disagreements will be resolved through discussion and conflicts resolved by a third reviewer (JD). We will contact study authors to resolve any uncertainties about extracted data.

#### Item 12. Data items

The following data items will be extracted:

1. Identification of the study
    a. Journal,
    b. Authors,
    c. Year,
    d. Citation,
    e. Research center/university/hospital/organization,
    f. Conflict of interest,
    g. Funding/sponsorship.
2. Methods
    a. Study aim,
    b. Study design,
    c. Participant demographics,
    d. Recruitment process,
    e. Inclusion,

f. Exclusion criteria,
g. Statistical analysis.

3. Main findings
   a. Exposure details,
   b. Diagnostic delays,
   c. Factors associated with diagnostic delay,
   d. Patients' experience,
   e. Relevant outcomes.

#### Item 13. Outcomes and prioritization

1. Primary outcome
   a. Diagnostic delay time (time from symptom onset to correct diagnosis) in people living with Myositis
2. Secondary outcomes
   a. Patient's experiences related to diagnostic delay
   b. Causes and consequences of diagnostic delay

#### Item 14. Quality assessment or risk of bias

The selected studies will be assessed for methodological quality or risk of bias using the Mixed Methods Appraisal Tool (MMAT) designed to critically appraise mixed method studies included in systematic reviews (Hong et al., 2018). Two independent review authors will conduct the quality appraisal. Any conflicts will be resolved with discussion and a third reviewer's vote (JD).

#### Item 15. Data synthesis

A systematic narrative synthesis will be undertaken to explore the findings of included studies in relation to time from symptom onset to diagnosis, and people's experiences related to delayed diagnosis in line with guidance from the Centre for Reviews and Dissemination (Centre for Reviews and Dissemination., 2009).

If extracted quantitative data are homogenous, a meta-analysis will be conducted using a random-effects model. Extracted qualitative data will be meta-synthesized using meta-aggregation. In line with meta-aggregation methods, findings (processed data) from qualitative studies will be extracted and aggregated into a single set of categories, which will then be further aggregated and synthesised into a set of statements that are meaningful for clinical practice.

Further methods and stages of meta-analysis will be discussed if collected data is quantitively synthesizable. The findings from the quantitative and qualitative studies will be reported separately; however, the discussion will be integrative of both.

#### Item 16. Confidence in cumulative estimate

If a meta-analysis is conducted, the quality/certainty of evidence for all quantitative outcomes will be judged using the Grading of Recommendations Assessment, Development and Evaluation (GRADE) working group methodology (Balshem et al., 2011). Certainty of the body of evidence will be assessed across the domains of risk of bias, consistency of effect, imprecision, indirectness, and publication bias. The certainty will be reported in four levels: high, moderate, low, and very low.

#### Item 17. Timeline and stages of review

Table 2. Timeline and process of systematic review

| | Started | Completed | Timeline |
|---|---|---|---|
| Protocol development | Yes | Yes | November, 2021 |
| Search strategy development | Yes | Yes | November 2021 |
| Preliminary literature search | Yes | Yes | November, 2021 |
| Literature search | Yes | No | November, 2021 |
| Piloting of the study selection process | No | No | January, 2021 |
| Study selection | No | No | January-February 2022 |
| Quality appraisal | No | No | February, 2022 |
| Data extraction | No | No | January- February 2022 |
| Data synthesis | No | No | February- March, 2022 |
| Writing paper | No | No | March-May, 2022 |

#### Version history

Table 3. Version history of systematic review protocol

| Date | Version number | Explanation |
|---|---|---|
| 28 October 2021 | Version 1.0 | First draft of review, “Diagnostic delay of Myositis: a protocol of an integrated systematic review” |
| 25 November 2021 | Version V2.0 | Second draft of review, “Diagnostic delay of Myositis: a protocol of an integrated systematic review” |

### Appendix 1.

Search terms used to develop final search string for PubMed search conducted on 9th[th] of December 2021.

#### Search string:

"myositis"[Title/Abstract] AND ("delay in diagnosis"[Title/Abstract] OR "diagnostic delay"[Title/Abstract] OR "misdiagnosis"[Title/Abstract] OR "time to diagnosis"[Title/Abstract] OR "incorrect diagnosis"[Title/Abstract] OR "missed diagnosis"[Title/Abstract] OR "delayed diagnosis"[Title/Abstract])

#### Search history

Table 4. Search history of Myositis search for PUBMED/MEDLINE

| Search number | Query | Search Details | Results |
|---|---|---|---|
| 11 | #1 AND #10 | "myositis"[Title/Abstract] AND ("delay in diagnosis"[Title/Abstract] OR "diagnostic delay"[Title/Abstract] OR "misdiagnosis"[Title/Abstract] OR "time to diagnosis"[Title/Abstract] OR "incorrect diagnosis"[Title/Abstract] OR "missed diagnosis"[Title/Abstract] OR "delayed diagnosis"[Title/Abstract] OR "slow diagnosis"[Title/Abstract]) | 96 |
| 10 | #2 OR #3 OR #4 OR #5 OR #6 OR #7 OR #8 OR #9 | "delay in diagnosis"[Title/Abstract] OR "diagnostic delay"[Title/Abstract] OR "misdiagnosis"[Title/Abstract] OR "time to diagnosis"[Title/Abstract] OR "incorrect diagnosis"[Title/Abstract] OR "missed diagnosis"[Title/Abstract] OR "delayed diagnosis"[Title/Abstract] | 38,185 |
| 9 | "slow diagnosis"[Title/Abstract] | "slow diagnosis"[Title/Abstract] | 8 |
| 8 | "delayed diagnosis"[Title/Abstract] | "delayed diagnosis"[Title/Abstract] | 8,589 |
| 7 | "missed diagnosis"[Title/Abstract] | "missed diagnosis"[Title/Abstract] | 2,399 |
| 6 | "incorrect diagnosis"[Title/Abstract] | "incorrect diagnosis"[Title/Abstract] | 1,299 |
| 5 | "time to diagnosis"[Title/Abstract] | "time to diagnosis"[Title/Abstract] | 2,661 |

| | | | |
|---|---|---|---|
| 4 | "misdiagnosis"[Title/Abstract] | "misdiagnosis"[Title/Abstract] | 16,610 |
| 3 | "diagnostic delay"[Title/Abstract] | "diagnostic delay"[Title/Abstract] | 3,203 |
| 2 | "delay in diagnosis"[Title/Abstract] | "delay in diagnosis"[Title/Abstract] | 5,967 |
| 1 | myositis[Title/Abstract] | "myositis"[Title/Abstract] | 11,085 |
